# Resistance to anti-orthopoxviral drug tecovirimat (TPOXX^®^) during the 2022 mpox outbreak in the US

**DOI:** 10.1101/2023.05.16.23289856

**Authors:** Todd G. Smith, Crystal M. Gigante, Nhien T. Wynn, Audrey Matheny, Whitni Davidson, Yong Yang, Rene Edgar Condori, Kyle O’Connell, Lynsey Kovar, Tracie L. Williams, Yon C. Yu, Brett W. Petersen, Nicolle Baird, David Lowe, Yu Li, Panayampalli S. Satheshkumar, Christina L. Hutson

## Abstract

**Background:** During the 2022 multinational outbreak of monkeypox virus (MPXV) clade IIb, the antiviral drug tecovirimat (TPOXX^®^) was deployed in the US on a large scale for the first time ever. The MPXV F13L gene homolog encodes the target of tecovirimat, and single amino acid changes in the F13 protein are known to cause resistance to tecovirimat in orthopoxviruses (OPXV).

**Methods:** Whole genome metagenomic sequencing and amplicon-based sequencing targeting the F13L gene was used to identify nine mutations previously reported to cause resistance in other OPXV along with ten novel mutations that have been identified from the 2022 mpox outbreak. A cytopathic effect assay, previously established at CDC as part of WHO smallpox research, was adapted to MPXV for tecovirimat phenotype testing of virus isolated from mpox patients.

**Results:** As of March 2023, in total, 70 isolates from 40 patients were tested, and 50 of these isolates from 26 patients were found to have a resistant phenotype. Most resistant isolates were associated with severely immunocompromised mpox patients on multiple courses of TPOXX treatment; while isolates with F13 mutations identified by routine surveillance of patients not treated with TPOXX have remained sensitive.

**Conclusions:** These data indicate that tecovirimat resistance is developing in immunocompromised patients treated with TPOXX and that for isolates that we have analyzed, the frequency of resistant viruses remain relatively low (< 1%) compared to the total number of patients treated with TPOXX. These findings inform our understanding of when tecovirimat resistance is likely to occur and highlight the need for additional OPXV therapeutics.

## Original Research

In May 2022, an outbreak of monkeypox virus (MPXV) clade IIb (formerly West African clade) was first identified in the US^1^. Since that time over 32,000 cases and 38 deaths associated with the outbreak have been identified in the US. Cases peaked the first week of August 2022 in the US with 459 cases per week. The US has identified more cases than any other country in the global outbreak^2^.

The FDA licensed the therapeutic agent TPOXX^®^ containing the drug tecovirimat (i.e. ST-246) under the animal rule for smallpox treatment in 2018^3^. Tecovirimat has been tested extensively in cell culture ^4-6^ and within many orthopoxvirus (OPXV) animal models^7-15^, including the non-human primate variola virus (VARV) model^16,17^. Although tecovirimat has shown efficacy against multiple OPXV, it has also been noted that nucleotide alterations to the orthopoxviral F13L gene homolog leading to amino acid (AA) substitutions in the F13 protein (also known as VP37) allows for resistance^4,18^. Additionally, resistance emerged during use of tecovirimat in an extended treatment course of an individual with progressive vaccinia^19^.

Because TPOXX is only licensed for treatment of smallpox, CDC holds an expanded access investigational new drug protocol for treatment of non-variola OPXV infections, including mpox. Since June 2022, 6929 patients have received TPOXX for mpox treatment in the US. A fraction of these have been severe cases where patients have moderate to severe immunocompromise usually due to uncontrolled HIV infection^20^. We have received specimens from 432 patients that received TPOXX and resistance was possible or suspected based on clinical data (Fig. 1A). Specimens have been tested from 40 patients, and a resistant phenotype has been confirmed in 26 patients (Fig 1B).

**Figure 1.**
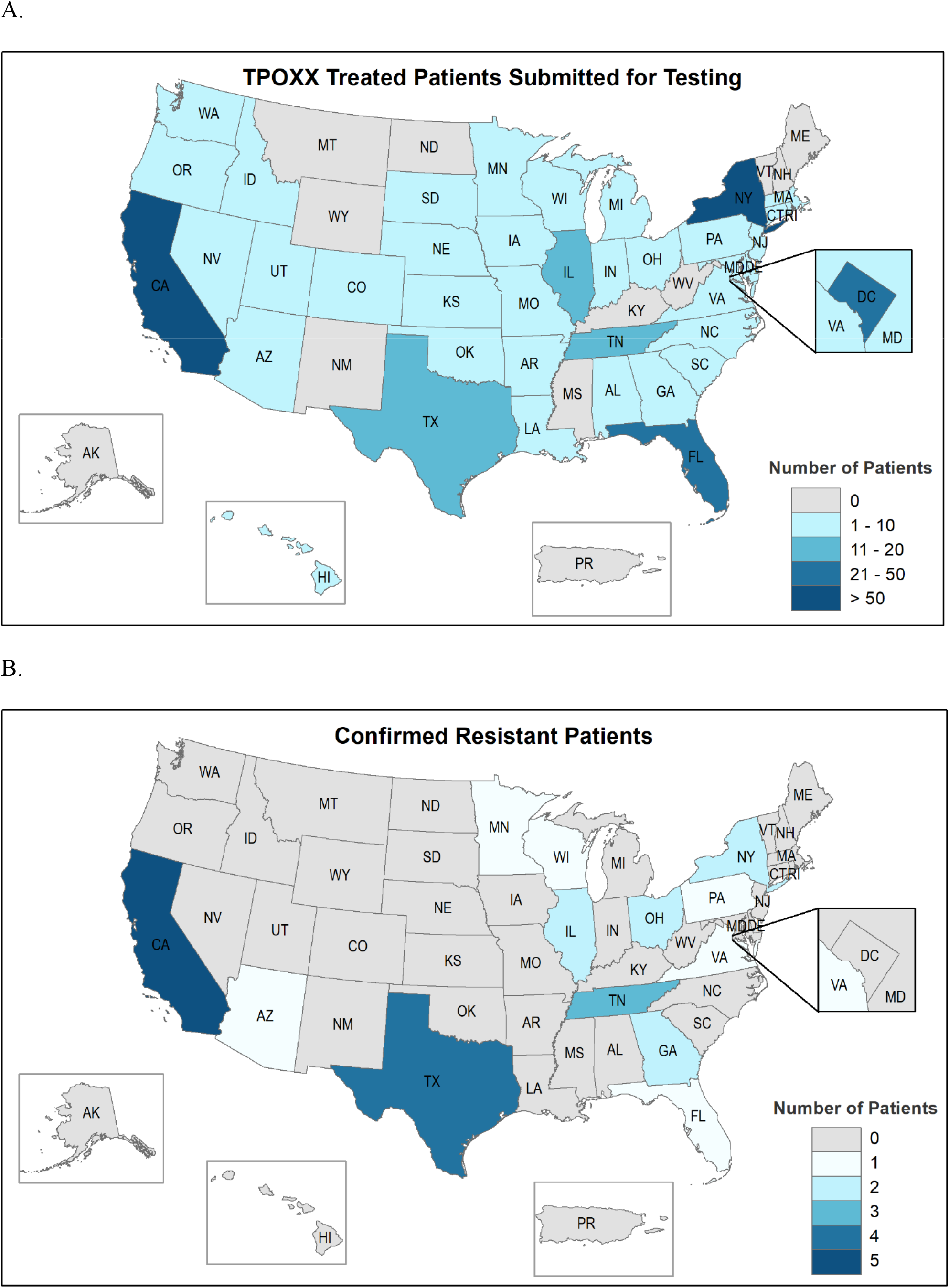
Surveillance for tecovirimat resistance in the US as of March, 2023

During the mpox outbreak, whole genome metagenomic sequencing and more recently amplicon-based sequencing targeting the F13L gene have been used to screen for changes in the MPXV F13L homolog (Table S1 including methods). A total of 2,181 CDC-generated sequences have been screened by either passive genomic surveillance (n=2,049) or targeted F13L sequencing (n=132). Only genomic sequencing completed at CDC was included because the raw data were required to find minor variants. The primary outbreak strain (MPXV clade IIb lineage B.1) has a substitution, E353K, in the F13 protein that is not present in the secondary outbreak strain (lineage A.2), historical clade IIb sequences from Nigeria, or MPXV clade IIa^21^. Since the E353K substitution was not previously described in other OPXV, the effect on tecovirimat phenotype was unknown. A cytopathic effect (CPE) assay, used at CDC to test VARV sensitivity to tecovirimat, was adapted to MPXV as described previously^21^. Briefly, MPXV was cultured from clinical specimens which were decoded but not anonymous. A gradient of tecovirimat was applied to confluent cell monolayers, then cells were infected with the isolated MPXV and incubated for 72 hours. Wells were fixed, stained, and absorbance at 570 nm was measured. The CPE assay was used to show that MPXV isolates with the E353K mutation remained sensitive to tecovirimat^21,22^.

In total, 86 samples from 55 patients produced sequences with AA changes (other than E353K) in the F13 protein relative to MPXV Clade IIb variant B.1 reference ON563414 (Table 1). Isolates with AA substitutions D100N, D217N, D248N and D256N, identified by routine sequencing of samples from patients not treated with TPOXX, have remained sensitive (Table 1). Nine AA mutations, A290V, A295E, D294V, H238Q, I372N, L297ins, N267D, N267del, and A288P, that were previously identified in other OPXV^18,19,23^, were confirmed resistant by phenotypic testing (Table 1). One confirmed resistance mutation, T289A, had not been described before the 2022 mpox outbreak^24^. T289A resulted in up to an 8-fold increase in the half-maximal effective concentration (EC_50_) when compared to the MPXV clade IIa reference strain (US, 2003). This position is part of the predicted tecovirimat binding site and adjacent to A288P and A290V which both confer resistance^18^. Five other AA substitutions (K174N, S215F, P243S, Y285H, R291K) were identified but the effect of these mutations has not yet been determined^24,25^. These mutations have only been observed and tested in combination with other known resistance mutations.

**Table 1.**
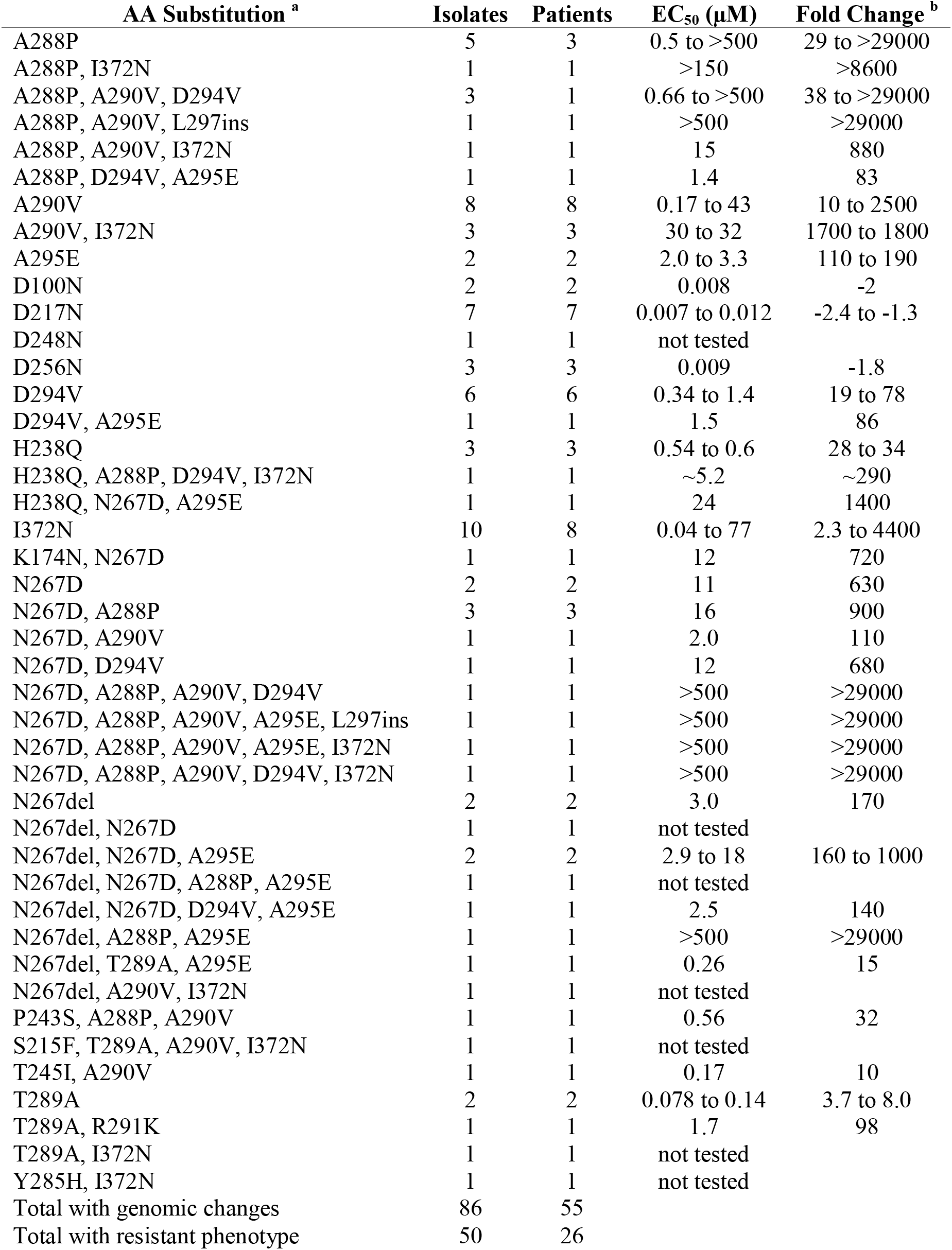
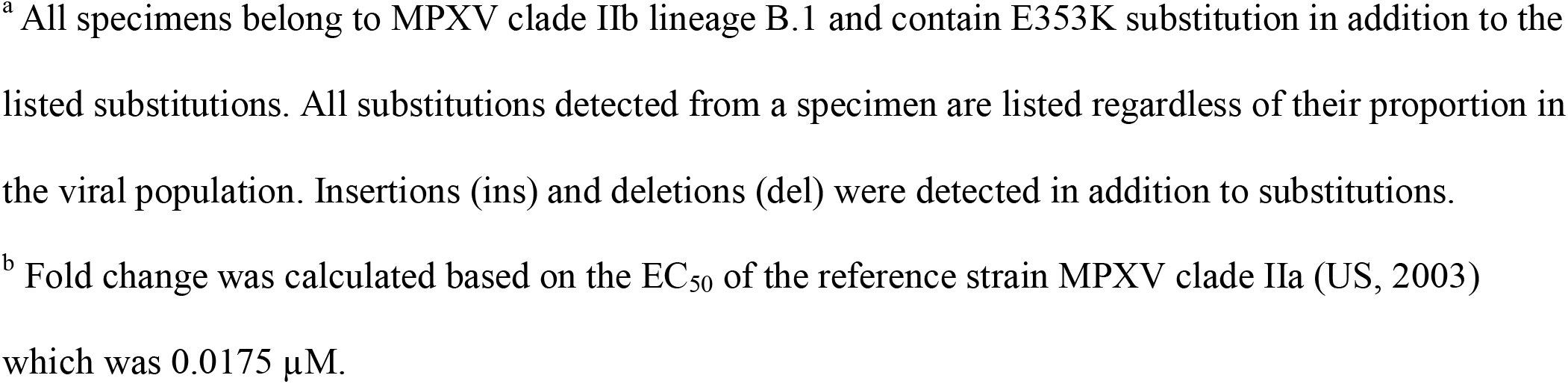
F13 mutations identified in patients (n=???) treated with TPOXX

| AA Substitution <sup>a</sup> | Isolates | Patients | EC<sub>50</sub> (μM) | Fold Change <sup>b</sup> |
| --- | --- | --- | --- | --- |
| A288P | 5 | 3 | 0.5 to >500 | 29 to >29000 |
| A288P, I372N | 1 | 1 | >150 | >8600 |
| A288P, A290V, D294V | 3 | 1 | 0.66 to >500 | 38 to >29000 |
| A288P, A290V, L297ins | 1 | 1 | >500 | >29000 |
| A288P, A290V, I372N | 1 | 1 | 15 | 880 |
| A288P, D294V, A295E | 1 | 1 | 1.4 | 83 |
| A290V | 8 | 8 | 0.17 to 43 | 10 to 2500 |
| A290V, I372N | 3 | 3 | 30 to 32 | 1700 to 1800 |
| A295E | 2 | 2 | 2.0 to 3.3 | 110 to 190 |
| D100N | 2 | 2 | 0.008 | -2 |
| D217N | 7 | 7 | 0.007 to 0.012 | -2.4 to -1.3 |
| D248N | 1 | 1 | not tested |  |
| D256N | 3 | 3 | 0.009 | -1.8 |
| D294V | 6 | 6 | 0.34 to 1.4 | 19 to 78 |
| D294V, A295E | 1 | 1 | 1.5 | 86 |
| H238Q | 3 | 3 | 0.54 to 0.6 | 28 to 34 |
| H238Q, A288P, D294V, I372N | 1 | 1 | ~5.2 | ~290 |
| H238Q, N267D, A295E | 1 | 1 | 24 | 1400 |
| I372N | 10 | 8 | 0.04 to 77 | 2.3 to 4400 |
| K174N, N267D | 1 | 1 | 12 | 720 |
| N267D | 2 | 2 | 11 | 630 |
| N267D, A288P | 3 | 3 | 16 | 900 |
| N267D, A290V | 1 | 1 | 2.0 | 110 |
| N267D, D294V | 1 | 1 | 12 | 680 |
| N267D, A288P, A290V, D294V | 1 | 1 | >500 | >29000 |
| N267D, A288P, A290V, A295E, L297ins | 1 | 1 | >500 | >29000 |
| N267D, A288P, A290V, A295E, I372N | 1 | 1 | >500 | >29000 |
| N267D, A288P, A290V, D294V, I372N | 1 | 1 | >500 | >29000 |
| N267del | 2 | 2 | 3.0 | 170 |
| N267del, N267D | 1 | 1 | not tested |  |
| N267del, N267D, A295E | 2 | 2 | 2.9 to 18 | 160 to 1000 |
| N267del, N267D, A288P, A295E | 1 | 1 | not tested |  |
| N267del, N267D, D294V, A295E | 1 | 1 | 2.5 | 140 |
| N267del, A288P, A295E | 1 | 1 | >500 | >29000 |
| N267del, T289A, A295E | 1 | 1 | 0.26 | 15 |
| N267del, A290V, I372N | 1 | 1 | not tested |  |
| P243S, A288P, A290V | 1 | 1 | 0.56 | 32 |
| S215F, T289A, A290V, I372N | 1 | 1 | not tested |  |
| T245I, A290V | 1 | 1 | 0.17 | 10 |
| T289A | 2 | 2 | 0.078 to 0.14 | 3.7 to 8.0 |
| T289A, R291K | 1 | 1 | 1.7 | 98 |
| T289A, I372N | 1 | 1 | not tested |  |
| Y285H, I372N | 1 | 1 | not tested |  |
| Total with genomic changes | 86 | 55 |  |  |
| Total with resistant phenotype | 50 | 26 |  |  |

<sup>a</sup> All specimens belong to MPXV clade IIb lineage B.1 and contain E353K substitution in addition to the listed substitutions. All substitutions detected from a specimen are listed regardless of their proportion in the viral population. Insertions (ins) and deletions (del) were detected in addition to substitutions.
<sup>b</sup> Fold change was calculated based on the EC<sub>50</sub> of the reference strain MPXV clade IIa (US, 2003) which was 0.0175 µM.

For phenotype testing, an isolate was considered resistant if the increase in the EC_50_ compared to the MPXV clade IIa (US, 2003) reference strain was ≥ 2-fold. Isolates with 2-to 9-fold change were considered partially resistant and isolates with ≥10-fold change were considered resistant^24^. Forty-three isolates from 22 patients were resistant, and seven isolates from five patients were partially resistant.

One patient with partially resistant isolates also had at least one other isolate that was resistant. The clinical relevance of partially resistant and/or resistant isolates remains unknown.

Multiple lines of evidence point to tecovirimat resistance developing during drug treatment in most patients. First, genome sequencing has revealed unique mutational profiles from different sample sites from the same patient (Fig. 2.A). Indicating different viral subpopulations were selected at different sites during treatment. Second, longitudinal sampling was investigated for four of the 26 patients with a resistant isolate and showed samples before tecovirimat treatment were sensitive whereas later samples were resistant (Fig. 2.B). An exception was found for one patient. T289A was detected in 58% of reads, with minor populations of A295E (9%) and N267del (22%), from a sample the day before the patient started tecovirimat treatment. A second sample from the same patient after tecovirimat treatment showed the T289A mutation alone was selected (93%). Additionally, N267del was detected in a cluster of cases with no known TPOXX treatment by Los Angeles County Department of Public Health (manuscript in preparation). Whether these drug-resistant cases were acquired from another person treated with tecovirimat is unknown but is a viable hypothesis. These rare cases show that viruses with mutations in F13L resulting in tecovirimat resistance can be transmitted from person to person.

**Figure 2.**
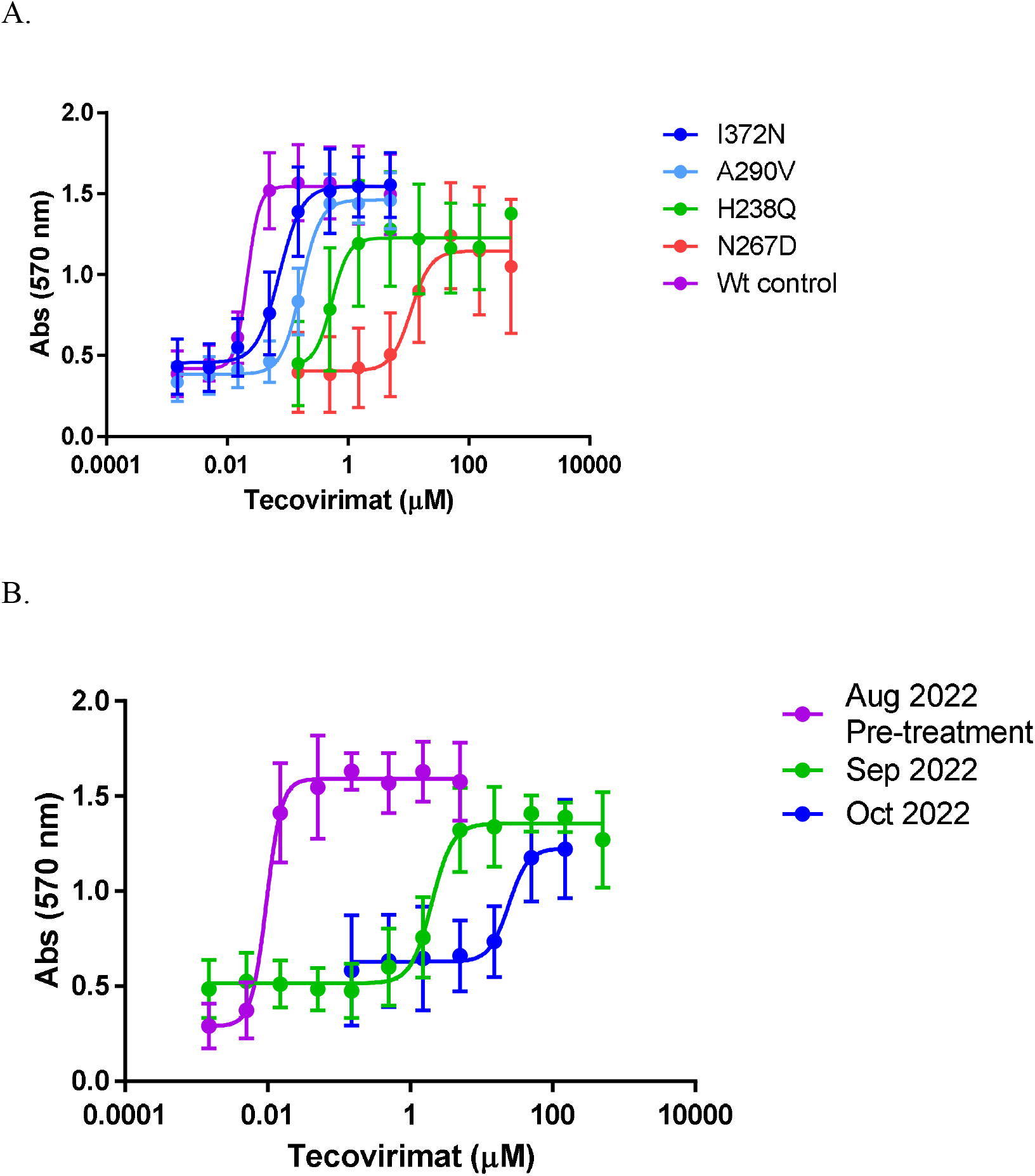
Examples of tecovirimat resistance Patient samples were sequenced, cultured, and subjected to tecovirimat sensitivity testing in a cytopathic effect assay. (A) Different samples from the same patient showed different F13 AA substitutions which result in different levels of resistance compared to the wild-type (Wt) control (MPXV clade IIa, US, 2003). (B) Samples from the same patient at different times before and after starting tecovirimat treatment in Aug 2022 showing sensitivity before drug treatment and increasing resistance after drug treatment.

For patients that had at least one specimen with confirmed tecovirimat resistance, all 26 had HIV infection. Of the 26 patients, 17 had a CD4^+^ T-cell count available, all 17 were below 350 cell/mm^3^, and 15 were below 200 cell/mm^3^. Five of the 26 patients died^24,25^. All five deceased patients had CD4^+^ T-cell counts below 200 cell/mm^3^. At least 23 of 26 patients were hospitalized. No medical history was available for one patient, and the medical history concerning hospitalization was not clear for the remaining two patients. All 26 patients received tecovirimat. Exact data on length of tecovirimat exposure is difficult to obtain because of possible noncompliance with oral administration and multiple rounds of treatment where drug administration stops and starts. We estimated the average length of tecovirimat treatment using the reported start date of TPOXX treatments. Dates were available for 21 of the 26 patients, and the average length of tecovirimat treatment was 32 days with a range of 14 to 77 days (standard regimen is 14 days).

Our report has several limitations. The phenotype assay is culture-based making it labor intensive and low throughput. As of March 2023, we have phenotyped70 specimens from 40 patients. The lag in testing means most of the specimens that have been tested are from September to November 2022, so results only give a retrospective sample of possible drug resistance. Submission of samples for tecovirimat sensitivity is voluntary (but encouraged for suspected tecovirimat resistance), so sampling bias may exist for certain physicians, hospitals, or states and may make it appear that certain states have more drug-resistance than others (Fig. 1). As genomic sequencing has increased, we have prioritized samples with predicted resistance mutations for phenotype testing. Mixed populations of cultured virus were tested due to the efficiency needed for a public health emergency. In the future, we will begin plaque purification for selected samples to test clonal populations.

Finally, these results confirm that tecovirimat resistance mutations are being selected in human mpox patients by tecovirimat treatment. Resistance has been confirmed in a small percentage of cases for which specimens have been sent to CDC, currently < 1% with the potential to be ∼5% as testing continues. Characteristics of patients with resistant isolates are very similar: HIV infection with very low CD4^+^ T-cell counts and potential for extensive tecovirimat exposure while hospitalized. In very rare cases, a drug-resistant virus appears to have been transmitted to another person. Genomic and phenotype testing are ongoing. These results may be useful when considering treatment for patients that match the clinical profile described within, and that aggressive early dosing/combination therapy regimens could be considered in those instances^20^. Results will also provide critical knowledge to potentially build a genomic assay for early detection of resistance mutations which could be used to inform clinical care decisions. This report describes the largest number of tecovirimat resistant isolates from humans reported to date and provides crucial data on the AA changes leading to resistance in MPXV paired with clinical outcomes. These combined data may inform decisions on tecovirimat utilization in the future. These findings also highlight the need for additional, well-tolerated OPXV therapeutics with different modes of action, particularly for use with immunocompromised patients.

For clinicians concerned about tecovirimat resistance, we encourage enrolling patients in the CDC VIRISMAP study (cdc.gov/poxvirus/mpox/clinicians/treatment.html) and the STOMP trial (stomptpoxx.org).

## Data Availability

All data produced in the present study are available upon reasonable request to the authors

## Ethics Statement

The activities in this report were reviewed by the Human Subjects Advisor in the National Center for Emerging and Zoonotic Diseases at the Centers for Disease Control and Prevention and determined that it does not meet the regulatory definition of research under provision 45 CFR 46.102(l); the activities fall under public health surveillance and do not require IRB review.

## Disclaimer

Use of trade names and commercial sources are for identification only and do not imply endorsement by the U.S. Department of Health and Human Services. The findings and conclusions in this report are those of the authors and do not necessarily represent the views of the Centers for Disease Control and Prevention or their institutions.

## Acknowledgements

This study did not receive any external funding. We thank Hui Zhao and members of CDC Division of Scientific Resources Genome Sequencing Lab, CDC Advanced Molecular Detection Biotechnology Core Facility Branch, and CDC Mpox Response Bioinformatics Unit for support with genome sequencing. We thank Yoshinori Nakazawa for technical assistance. We thank all clinicians, hospitals, state and local public health laboratories (PHLs) that have submitted mpox specimens to CDC. We thank the following partners that have specifically submitted mpox specimens for tecovirimat resistance testing at the time of this report: California -Los Angeles County Public Health Laboratory; California Department of Public Health, Microbial Diseases Laboratory; California Department of Public Health, Viral and Rickettsial Disease Laboratory; Florida Bureau of Public Health Laboratories - Jacksonville; Florida Bureau of Public Health Laboratories - Miami; Florida Bureau of Public Health Laboratories - Tampa; Georgia Department of Public Health Laboratory; Illinois Department of Public Health, Chicago Laboratory; Indiana State Department of Health Laboratory Services; Louisiana Office of Public Health Laboratories; Maryland Department of Health, Laboratories Administration; North Carolina State Laboratory of Public Health; Nevada State Public Health Laboratory; New York City Department of Health and Mental Hygiene; Ohio Department of Health Laboratory; Pennsylvania Department of Health, Bureau of Laboratories; Rhode Island State Health Laboratories; Tennessee Division of Laboratory Services; Texas Department of State Health Services, Lab Services Section; Texas-Houston Health Department Laboratory; Virginia Division of Consolidated Laboratory Services.

