## Supplemental Material for "Resistance to anti-orthopoxviral drug tecovirimat (TPOXX^®^) during the 2022 mpox outbreak in the US"

**Supplemental Methods**

DNA was extracted from lesion swabs using EZ-1 DNA tissue kit (Qiagen) followed by heat inactivation at 56°C for ≥ 1 hour. Monkeypox virus infection was confirmed by real-time PCR using a cladeII-specific monkeypox virus real-time PCR assay as described in Li et al. (2010).

F13L amplicon sequencing: 5 µL of MPXV DNA was used as input to the primary PCR reaction with tagged primers (F13L Forward: ont_tag-GACCTTCTTCATTTCGTGCCA, F13L: Reverse ont_tag-AATGTGGCCATTTGCATCGG), where ont_tag was added as described by the manufacturer (Oxford Nanopore Technologies, EXP-PBC096). Reaction contained 10 µL 2x GC Buffer I (Takara, RR02AG), 0.2 µL long amplicon Taq polymerase (Takara, RR02AG), 0.5 µL each of F13L forward and reverse primers at 20 µM, 2 µL dNTPs (Takara, RR02AG), and 1.8 µL nuclease-free water. PCR reaction was run for 2 minutes at 94°C followed by 25 to 35 cycles of 30 s at 94°C, 30 s at 56°C, and 1 minute at 72°C, followed by 5 minutes at 72°C. Cycle number was determined by Clade II-specific Ct value: 25 cycles for Ct 20 – 25, 30 times for Ct 25 – 30, and 35 cycles for Ct >30. Samples with Ct < 20 were diluted 100-fold then run for 25 cycles. PCR reactions were cleaned up with 0.65x AMPure XP beads (Beckman). Barcoding PCR was performed according to the manufacturer’s instructions (Oxford Nanopore Technologies, EXP-PBC096) using 20 µL reactions, Takara LA taq with GC buffers as above (Takara, RR02AG), 1 minute extension time and 12 cycles of PCR. PCR reactions were cleaned up with 0.65x AMPure XP beads (Beckman). Samples concentrations were estimated using a Qubit and pooled at equal concentrations. Library preparation was performed using the SQK-LSK109 kit for sequencing on the Flongle device, according to the manufacturer (Oxford Nanopore Technologies). Basecalling was performed using guppy 6.1.2 (Oxford Nanopore Technologies) and flags –kit SQK-LSK109 –flowcell FLO-FLG001 –barcode_kits EXP-PBC096 –trim_barcodes –require_barcodes_both_ends. Nanopore reads were trimmed to remove 55 bp from each end (seqtk 1.0, https://github.com/lh3/seqtk) and all reads below 50 bp were removed (trimmomatic 0.39, https://github.com/timflutre/trimmomatic) before mapping to MPXV Nigeria reference MT903344 with 6,000 bp removed from the left terminus using minimap2 2.16 (https://github.com/lh3/minimap2). Variants were called using ivar 1.3.1 (<https://andersen-lab.github.io/ivar/html/manualpage.html>hid) and samtools 1.7 (<https://github.com/samtools/samtools>) using the command samtools mpileup -aa -A -B -Q 0 -d 80000 mappingfile.bam | ivar variants -p samplename -t 0.05 -m 5 -q 20.

Illumina metagenomics sequencing

Extracted DNA (15 µL) was used as input for the Illumina DNA Prep method according to the manufacturer’s protocol except one half reagent volumes were used throughout. Libraries were visualized using an Agilent Fragment Analyzer instrument and an HS NGS Fragment Kit (Agilent Technologies Inc., Santa Clara, CA). Forty-eight samples were pooled at approximately equal molarity generating 200 pM final loading concentration and sequenced on an Illumina NovaSeq 6000 instrument using the 300 cycle SP sequencing components. Orthopoxvirus reads were filtered using Kraken2 v2.1.2 (Wood et al., 2019) run with default settings, using a database that included human genome for negative selection and MPXV genomes for positive selection. We used seqtk v1.3 `subseq` (Li, 2012) to subsample our reads to orthopoxvirus with default settings and the ‘---no-name’ flag, then used fastp v0.23.2 (Chen et al., 2018) with to trim and clean our filtered reads. Reads were aligned to MPXV Clade IIb reference genome (UK-P2; MT903344.1) using bwa mem v0.7.17 (Li, 2013) then sorted using Samtools v.1.15.1 (Li et al., 2009). F13L variants were called using iVar v.1.3.1 (Castellano et al., 2021) with the following parameters: samtools mpileup -aa -A -d 600000 -B -Q 0 $PREFIX.BAM | ivar variants -p $PREFIX -r $REFERENCE -q 20 -t 0.05 -m 5. We converted from tsv to vcf format using a custom python script (https://github.com/jts/ncov-tools/blob/master/workflow/scripts/ivar_variants_to_vcf.py), filtering for an allele frequency of 0.05 and > 5 supporting reads. For both ONT and Illumina data, only variants with allele frequency >10% are reported here.

**Table S1.** F13 mutations identified in 15 U.S. mpox cases from 2022.

| **Case** | **Specimen** | **Mutation** | **Illumina** | **ONT** | **Difference** | **Illumina Depth** | **ONT Depth** |
| --- | --- | --- | --- | --- | --- | --- | --- |
| 1 | 1 | N267D | 100.00% | 98.90% | 1.10% | 37 | 366 |
| 2 | 1 | A288P | 26.90% | 29.30% | -2.40% | 2831 | 1617 |
|  |  | D294V | 26.10% | 18.00% | 8.10% |  |  |
|  |  | D301del | 23.60% | 17.10% | 6.50% |  |  |
| 2 | 2 | A288P | 18.59% | 19.41% | -0.82% | 1926 | 4670 |
|  |  | A290V | 26.88% | 26.50% | 0.38% |  |  |
|  |  | D294V | 24.80% | 16.64% | 8.16% |  |  |
| 2 | 3 | A288P | 13.97% | 14.78% | -0.81% | 1446 | 5723 |
|  |  | A290V | 20.61% | 22.44% | -1.83% |  |  |
|  |  | L297ins | 37.59% | 33.22% | 4.36% |  |  |
| 3 | 1 | D294V | 100.00% | 96.60% | 3.40% | 75 | 70 |
| 4 | 1 | T289A | 100.00% | 100.00% | 0.00% | 71 | 162 |
| 5 | 1 | D294V | 99.20% | 98.20% | 1.00% | 118 | 148 |
| 6 | 1 | N267del | 22.20% | 20.00% | 2.20% | 90 | 5962 |
|  |  | T289A | 58.20% | 61.00% | -2.80% |  |  |
|  |  | A295E | 9.46% | 11.20% | -1.74% |  |  |
| 6 | 2 | T289A | 92.90% | 89.90% | 3.00% | 114 | 4461 |
|  |  | R291K | 30.70% | 34.10% | -3.40% |  |  |
| 7 | 1 | N267D | 59.11% | 66.59% | -7.48% | 291 | 317 |
|  |  | D294V | 29.51% | 17.02% | 12.49% |  |  |
| 7 | 2 | N267del | 89.81% | 72.85% | 16.97% | 373 | 1563 |
| 8 | 1 | A295E | 100.00% | 98.52% | 1.48% | 37 | 2456 |
| 8 | 2 | N267del | 47.79% | 30.68% | 17.11% | 66 | 138 |
|  |  | A288P | 22.06% | 28.22% | -6.16% |  |  |
|  |  | A295E | 15.15% | 27.11% | -11.96% |  |  |
| 9 | 1 | N267D | 25.42% | 29.78% | -4.35% | 118 | 131 |
|  |  | A288P | ND | 11.76% | missed |  |  |
|  |  | A290V | 16.24% | 12.00% | 4.24% |  |  |
|  |  | A295E | 21.01% | 11.61% | 9.40% |  |  |
|  |  | I372N | 17.53% | 15.26% | 2.26% |  |  |
| 10 | 1 | A290V | 95.00% | 84.29% | 10.71% | 20 | 5377 |
|  |  | T245I | ND | 11.75% | missed |  |  |
| 11 | 1 | A290V | 100.00% | 97.80% | 2.20% | 161 | 3373 |
| 11 | 2 | I372N | 100.00% | 85.72% | 14.28% | 334 | 5176 |
| 12 | 1 | N267D | 18.32% | 28.07% | -9.75% | 198 | 418 |
|  |  | A295E | 60.32% | 56.01% | 4.31% |  |  |
| 12 | 2 | N267del | 78.40% | 67.94% | 10.46% | 125 | 4824 |
| 13 | 1 | A288P | 54.94% | 53.75% | 1.19% | 134 | 1241 |
|  |  | N267D | 43.28% | 48.72% | -5.44% |  |  |
| 14 | 1 | Y285H | 10.13% | 8.83% | 1.30% | 60 | 4092 |
|  |  | I372N | 90.00% | 77.04% | 12.96% |  |  |
| 15 | 1 | D217N | 100.00% | 98.97% | 1.03% | 19 | 91 |

DNA extracted from each specimen was sequenced by direct DNA sequencing on an Illumina NovaSeq 6000 or targeted F13L amplicon sequencing on an Oxford Nanopore MinIon (see supplemental methods). Percent of reads with each mutation is shown for the two methods. Two minor alleles were not detected by the direct DNA sequencing method (ND not detected). Allele frequencies less than 10% were not reported unless it was detected at >10% by the other method. Average read depth is included. For some cases, multiple specimens collected from different anatomical sites yielded different mutational patterns. Amino acid deletion (del) and insertion (ins) mutations are included.
